# Early life serological profiles and the development of natural protective humoral immunity to *Streptococcus pyogenes* in a high burden setting

**DOI:** 10.1101/2025.02.11.25322090

**Authors:** Alexander J Keeley, Fatoumata E Camara, Edwin Armitage, Gabrielle de Crombrugghe, Jainaba Sillah, Modou Lamin Fofana, Victoria Rollinson, Elina Senghore, Musukoi Jammeh, Alana L Whitcombe, Amat Bittaye, Haddy Ceesay, Isatou Ceesay, Bunja Samateh, Muhammed Manneh, Martina Carducci, Luca Rovetini, Elena Boero, Luisa Massai, Chilel Sanyang, Ousman Camara, Ebrima Cessay, Miren Iturriza, Danilo Moriel Gomes, Adam Kucharski, Pierre R Smeesters, Anne Botteaux, Ya Jankey Jayne, Nicole J Moreland, Ed Clarke, Beate Kampmann, Michael Marks, Omar Rossi, Henrik Salje, Claire E Turner, Thushan I de Silva

## Abstract

*Streptococcus pyogenes* leads to 500,000 deaths annually; many due to rheumatic heart disease in low-income settings. Limited understanding of natural protective immunity to *S. pyogenes* hinders vaccine development. We describe the evolution of serological profiles to conserved vaccine*-*antigens and type-specific M peptides from birth and throughout the life course in The Gambia. As placentally-transferred IgG waned after birth, serological evidence of new exposure was seen in 23% infants during the first year of life. Following culture-confirmed *S. pyogenes* events, the highest IgG increases occurred in children under two years following both pharyngeal and skin disease, and asymptomatic carriage at both sites. Higher IgG to conserved antigens SLO, SpyCEP and SpyAD correlated with functional activity and were associated with protection from culture-confirmed events following adjustment for age and anti-M protein IgG levels. Our data provide the first evidence of protection associated with humoral immunity to conserved vaccine candidate antigens in humans.

## Introduction

*Streptococcus pyogenes* (Group A *Streptococcus*) is a major global pathogen responsible for 500,000 deaths annually, with the majority due to Rheumatic Heart Disease (RHD) caused by long term pathological immune sequelae.^1,2^ An effective and equitable *S. pyogenes* vaccine is a global priority,^3^ yet few candidates are currently in clinical development.^3,4^ In addition to preventing invasive infections, a *S. pyogenes* vaccine needs to prevent throat and skin (pyoderma) infections, and perhaps asymptomatic carriage in children, that lead to pathological immune priming responsible for RHD.^5^ Much of the *S. pyogenes* disease burden, including RHD, is experienced in low- and middle-income countries (LMIC), where skin infections are more common than in high-income countries (HIC).^1,2,6^

A recognised scientific barrier to developing a *S. pyogenes* vaccine is the lack of understanding of naturally occurring immunity, particularly to protect against pharyngitis and pyoderma, which represent endpoints for future vaccine trials.^7,8^ The prevalence of *S. pyogenes* pharyngitis and pyoderma progressively reduce from childhood to adulthood, suggesting that naturally protective immunity is acquired through repeated exposures.^9,10^ Moreover, pooled intravenous immunoglobulin (IVIG) can promote opsonization, phagocytosis, and killing of bacteria *in vitro.*^11,12^ Together, these findings suggest that naturally occurring humoral immunity to *S. pyogenes* is one mechanism protecting adults from infection.

*S. pyogenes* vaccine development has taken two broad approaches. Initial efforts focused on the M protein, a major surface-expressed virulence factor encoded by the *emm* gene, also used for strain typing.^3,13,14^ The M protein is immunogenic, with type-specific antibodies shown to protect in animal models and in limited observational human studies.^15–19^ With over 275 *emm* types, multivalent M protein vaccines are limited by the high *emm*/M type diversity in LMIC.^20,21^ An alternative approach focuses on conserved antigens such as the *S. pyogenes* cell envelope protease (SpyCEP), *S. pyogenes* adhesion and division protein (SpyAD), Streptolysin O (SLO) and the Group A *Streptococcus* carbohydrate (GAC).^26^ While animal models and genomic analyses demonstrate a promising role for these as vaccine antigens, their contribution to natural protection in humans remains unknown.^26–31^ With both multivalent M-protein and conserved antigen vaccines in development, understanding evolution of natural immunity to these different antigens in early life and their relative roles in protection remains vital.

Within prospective observational cohort studies in The Gambia spanning the entire life course, we characterize serological profiles to leading conserved *S. pyogenes* vaccine antigens and *emm* type-specific M protein hypervariable regions (HVRs). We describe the age-, carriage-, and disease-related changes in antibody levels, and for the first time demonstrate a surrogate of protection against culture-confirmed *S. pyogenes* events associated with antibodies to conserved vaccine antigens.

## Results

### Study cohorts and S. pyogenes events

The dynamics of *S. pyogenes*-specific antibodies, carriage and disease events were measured in a prospective, household cohort study conducted in the urban area Sukuta, The Gambia, over a 13-month period in 2021-2022 (SpyCATS, NCT05117528).^10^ 442 individuals from 44 households were recruited and followed up at monthly visits; comprising 256 children <18 years (58%) and a median age of 15 (range 0-85, IQR 6-10), 233 (58%) female participants, and a median household size of seven (IQR 6-10). Participants were also seen between monthly visits if new symptoms consistent with pyoderma or pharyngitis were reported. Incidence and prevalence of *S. pyogenes* in this cohort have been reported previously.^10^ 108 *S. pyogenes* disease events (16 pharyngitis, 91 pyoderma, 1 mixed) and 90 carriage events (49 pharyngeal, 41 skin) were identified by bacterial culture in 141 individuals during the study (Fig S1). For greater resolution of antibody dynamics during the first year of life, serum samples from 94 mother-child pairs recruited to a clinical trial of meningococcal conjugate vaccine in pregnancy in 2018-2019 at an urban clinic in The Gambia (NCT03746665) were additionally included. The median age of mothers was 26 (IQR 23-29), with 35 (37%) female children. Fig 1 provides an overview of the study design.

**Figure 1:**
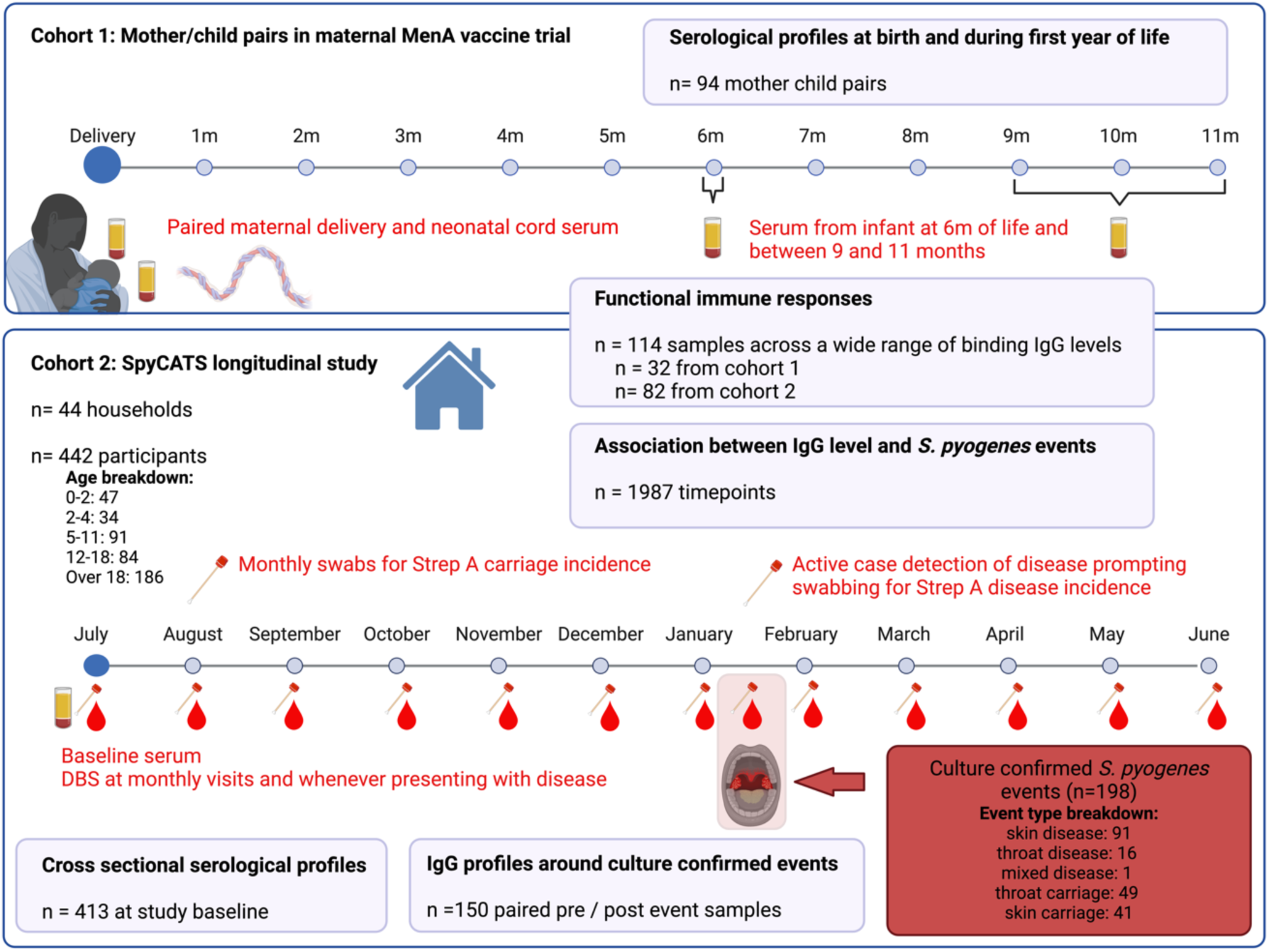
Study design and participants. Cohort 1 consisted of 94 mother child pairs from The Gambia recruited to a maternal vaccination trial with meningococcal conjugate vaccine. The newborn infants were followed through the first year of life. Cohort 2 were participants in the *Streptococcus pyogenes* carriage acquisition, persistence and transmission dynamics within households in The Gambia (SpyCATS) household cohort study. Red text indicates the sampling framework within both cohorts. In the SpyCATS study participants were swabbed from normal throat and skin to detect carriage. Participants could report disease symptoms (sore throat and skin sores) to the study team prompting swabbing from the relevant site to detect disease events. Antibodies were measured from serum collected at study baseline and from dried blood spot (DBS) at monthly visits and at any disease presentation. Purple boxes represent the number of samples included in each analysis. Breakdown of age groups and event types from the SpyCATS study is provided. Created in BioRender.com

### Waning of maternal IgG to conserved S. pyogenes vaccine antigens is followed by a rapid increase in early years of life

IgG specific to the conserved vaccine antigens GAC, SLO, SpyAD, and SpyCEP were measured from mother-child pairs in paired neonatal-cord and maternal-serum samples at delivery and in serum during the first year of life, along with DNAseB antibodies which are additionally used as serological evidence of *S. pyogenes* infection^32,33^. Antigen-specific antibody levels were quantified against an IVIG standard curve and expressed as Relative Luminex Units (RLU)/mL. Efficient placental transfer of *S. pyogenes*-specific IgG was observed, with no difference between paired maternal and cord sera at delivery in levels of SLO, SpyAD, SpyCEP and DNAseB (p>0.1 for all; Fig 2A & B). GAC-specific IgG was lower in cord sera compared to maternal samples, (5.08 vs 5.25 log10 RLU/mL, p<0.0001; Fig 2A & B). Waning of *S. pyogenes* antigen-specific IgG was observed in most children during the first 11 months of life (Fig 2A). Between six months and the subsequent sample (9,10, or 11 months), 22 infants (23%) demonstrated serological evidence of new *S. pyogenes* exposure, defined as any increase in IgG level to ≥2 antigens or >0.5 log10 RLU/mL increase to a single antigen (Fig 2A). The magnitude of antibody boosting to individual antigens was variable, with only two infants demonstrating IgG rises to all five antigens (Fig 2C). IgG dynamics across the life course to GAC, SLO, SpyAD, SpyCEP and DNAseB were explored using serum at recruitment to the SpyCATS study to model age-stratified antibody distributions (n=413, age range 0-85 years). IgG levels rose rapidly during early childhood with a plateau observed for all antigens by five years, and waning seen in older age in SLO-, SpyAD- and DNAseB-specific IgG (Fig 2D). Strong positive correlations were seen between all five antibody responses, with coefficients ranging from 0.65 to 0.86 (p<0.0001 for all comparisons, Fig S2).

**Figure 2:**
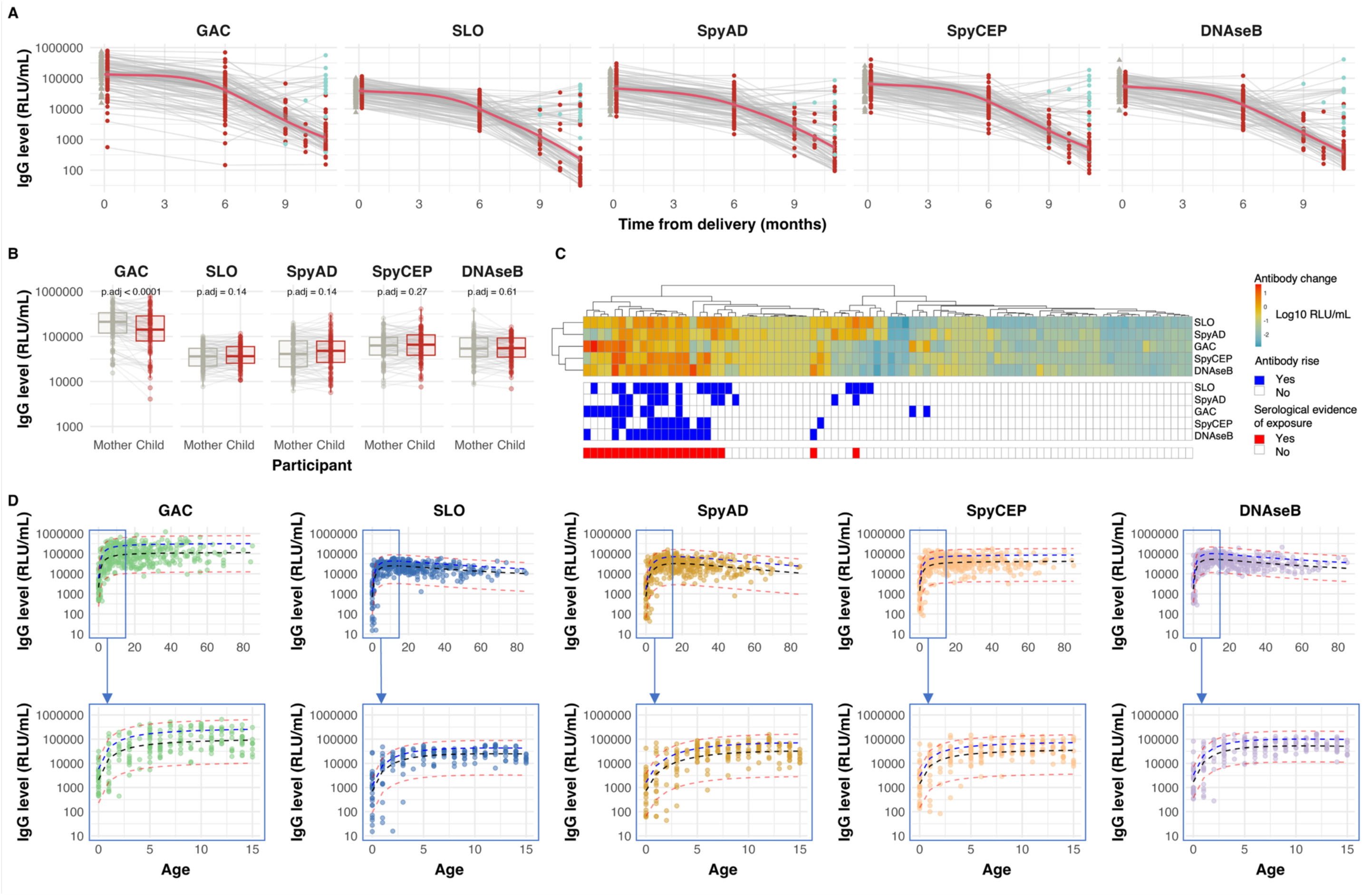
Antibody profiles to conserved S. pyogenes antigens over the life course. (**A**) Longitudinal antibody profiles in the first year of life from mother-child pairs (n=94). IgG titres in blood are expressed as Relative Luminex Units per mL (RLU/mL). Grey triangles represent maternal delivery samples. Dots represent neonatal samples. Red dots indicate neonatal samples where no titre increase between 6 months and subsequent visits was observed. Red line shows mean titre, with grey shading representing the 95% confidence interval (loess method used for generating waning profile). Green dots indicate neonatal samples with serological evidence of exposure, defined as increase in IgG level to two or more antigens or >0.5 log10 RLU/mL increase to a single antigen. (B) IgG levels in n=94 paired maternal serum and neonatal cord blood at delivery. IgG levels were compared using a paired Wilcoxon signed-rank test. P-values were adjusted for multiple testing using the false discovery rate (FDR) correction. (C) Heterogeneity of serological responses in early life from n=86 participants within the first year of life whose IgG level could be quantified to all 5 antigens at 6 months and one subsequent visit between 9 and 11 months. Each column represents an individual participant. Top panel demonstrates the magnitude of absolute change in log10 transformed IgG antibody levels between the 6-month visit and a subsequent visit. Dendrograms represent hierarchical clustering that was performed to test for similarities in antibody reactivity between individual participants (columns) and between antigens (rows) using Euclidean distance as the distance measure and complete linkage as the clustering method. Middle panel indicates binary responses to individual antigens in each individual between 6m visit and the subsequent visit: blue for positive changes (increase in antibody level) and white for negative changes (decrease in antibody level). Bottom panel indicates participants with serological evidence of exposure defined as an increase in IgG levels to two or more antigens, or to one antigen of at least 0.5 log10 RLU/mL. (D) Cross-sectional IgG blood profiles by age. Top panel shows entire age range (0-85 years) and bottom panel shows titres in children aged 0-15 years. In age plots, dotted lines represent the median (black), 80th centile (blue), and 2.5^th^ and 97th centiles (red), as calculated using a fractional polynomial model to predict titres by age.

### Disease and carriage events increase conserved S. pyogenes antigen-specific IgG, with the greatest rises seen in infancy

The kinetics of IgG levels to GAC, SLO, SpyAD, SpyCEP, and DNAseB before and after culture-confirmed *S. pyogenes* events were explored using serially collected dried blood spots (DBS). Paired pre- and post-event DBS samples were available for 150 events (13 pharyngitis, 67 pyoderma, 1 mixed disease, 34 pharyngeal carriage, 35 skin carriage). The median time between samples was 70 days (IQR 59-95). Significant rises in IgG levels to all five antigens were seen following events, ranging from 0.10 to 0.19 log10 RLU/mL (p<0.0001 for all comparisons, Fig S3A), although with substantial heterogeneity. Younger participants, particularly those under two years, had higher IgG rises (Fig 3A), with responses following all event types (Fig S3B&C). In a generalized linear mixed-effects model accounting for age and event type, participants under two years had significantly greater absolute increases in IgG levels, ranging from 0.25 to 0.54 log10 RLU/mL compared to adults (GAC p=0.011, SLO p=0.00017, SpyAD p=0.0068, SpyCEP p=0.00013, DNAseB p=0.013, Fig 3B). Baseline IgG levels and absolute increases following events were inversely correlated (coefficients -0.5 to -0.83, p<0.0001 for all antigens, Fig 3C). Rises in IgG were equivalent following pyoderma, and asymptomatic pharyngeal and skin carriage, when compared to pharyngitis (Fig 3C). In individuals with no culture-confirmed *S. pyogenes* during the study, IgG levels remained stable over the 13-month period, other than in infants under two years where several individuals had IgG rises and others demonstrated waning from baseline levels (Fig 3D).

**Figure 3:**
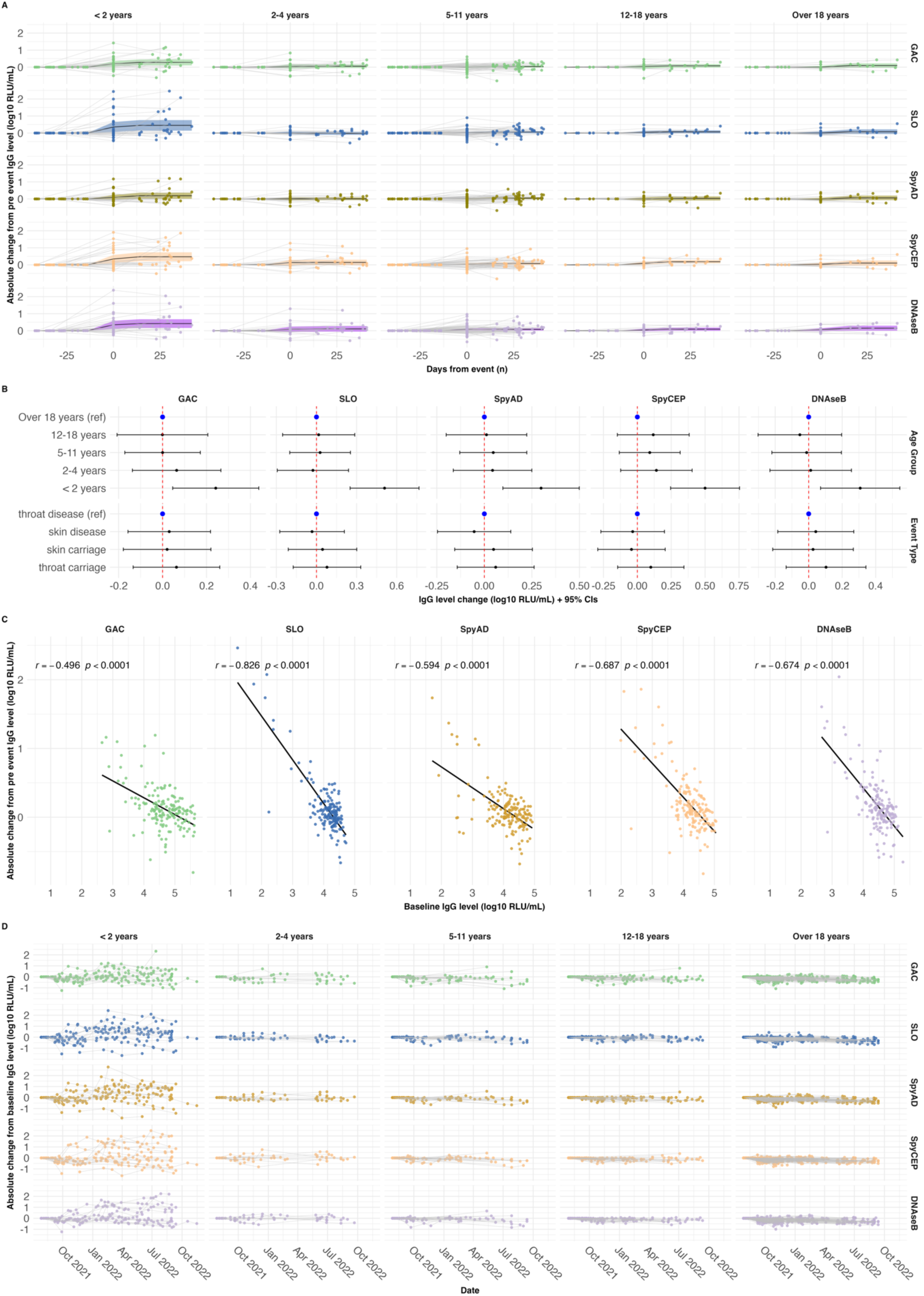
Blood IgG antibody profiles around culture-confirmed S. pyogenes events. (A) Individual IgG antibody profiles by age group around microbiologically confirmed *S. pyoge*nes events, where pre-event titres and at least one subsequent titre were measured (n=163 events). IgG was normalized to pre-event levels. Each dot represents an individual IgG level relative to the baseline titre. Grey lines connect individual participants’ IgG measured before, during and after events. Solid black lines represent the geometric mean log10 transformed IgG level changes across participants, grouped by temporal relationship to the event. Shaded areas around the lines represent 95% confidence intervals(B): Association between baseline (pre-event) IgG level and absolute increase in IgG level between pre and post event. The association between pre-event IgG level and absolute changes in titres was assessed using Pearson’s coefficient (C) Forest plot showing the association between age group and event type with absolute IgG level changes around Strep A events. The plot shows estimated differences between groups with 95% confidence intervals, calculated from mixed-effects linear regression models. (D) Longitudinal blood IgG profiles in participants (n=290) without microbiologically confirmed Strep A events during the study period. IgG levels were normalized to individual participants’ baseline titres. Each dot represents an individual antibody titre relative to baseline, and grey lines connect titres measured over time. RLU = Relative Luminex Unit.

### Higher IgG levels to SpyCEP, SpyAD and SLO are associated with a reduced risk of S. pyogenes events

IgG to conserved *S. pyogenes* vaccine antigens from 1987 timepoints in 431 participants were used to explore protection against 196 culture-confirmed events (Fig S1B). This included measurements at baseline, before, during, and after events in cases, and before and after events in household contacts where no *S. pyogenes* culture-confirmed event was detected at the time of an index event. At each IgG threshold, the proportion of visits with *S. pyogenes* events in the subsequent 45 days was determined (Fig S4A), and mixed-effects logistic regression models used to establish the association between *S. pyogenes*-specific IgG levels and the probability of subsequent *S. pyogenes* events. With this model, only higher anti-SpyCEP IgG was associated with reduced odds of *S. pyogenes* events (odds ratio (OR) 0.68, 95% confidence intervals (CI) 0.5-0.92, p=0.012; Fig S4B). For vaccine antigens SLO, SpyAD and SpyCEP, but not GAC nor DNAseB, the relationship between IgG levels and probability of events appeared non-linear, characterized by a plateau effect at lower antibody levels, followed by a downward slope (Fig S4, Fig 4A). We therefore employed piecewise regression to model the distinct portions of the relationship for SLO, SpyAD and SpyCEP. Transition points between portions were determined visually (Fig S4) and confirmed with iterative point increments and assessment of AIC, where values within 2 were considered comparable. Above the transition point, a significant reduction in culture-confirmed events within 45 days was seen for SLO (OR 0.06, CI 0.01-0.49, p=0.008), SpyAD (OR 0.34, CI 0.15-0.77, p=0.009) and SpyCEP (OR 0.25, 0.09-0.68, p=0.006, Fig 4B). IgG levels to all three antigens remained associated with protection in models adjusting for age, sex, and household size (Fig 4D).

**Figure 4:**
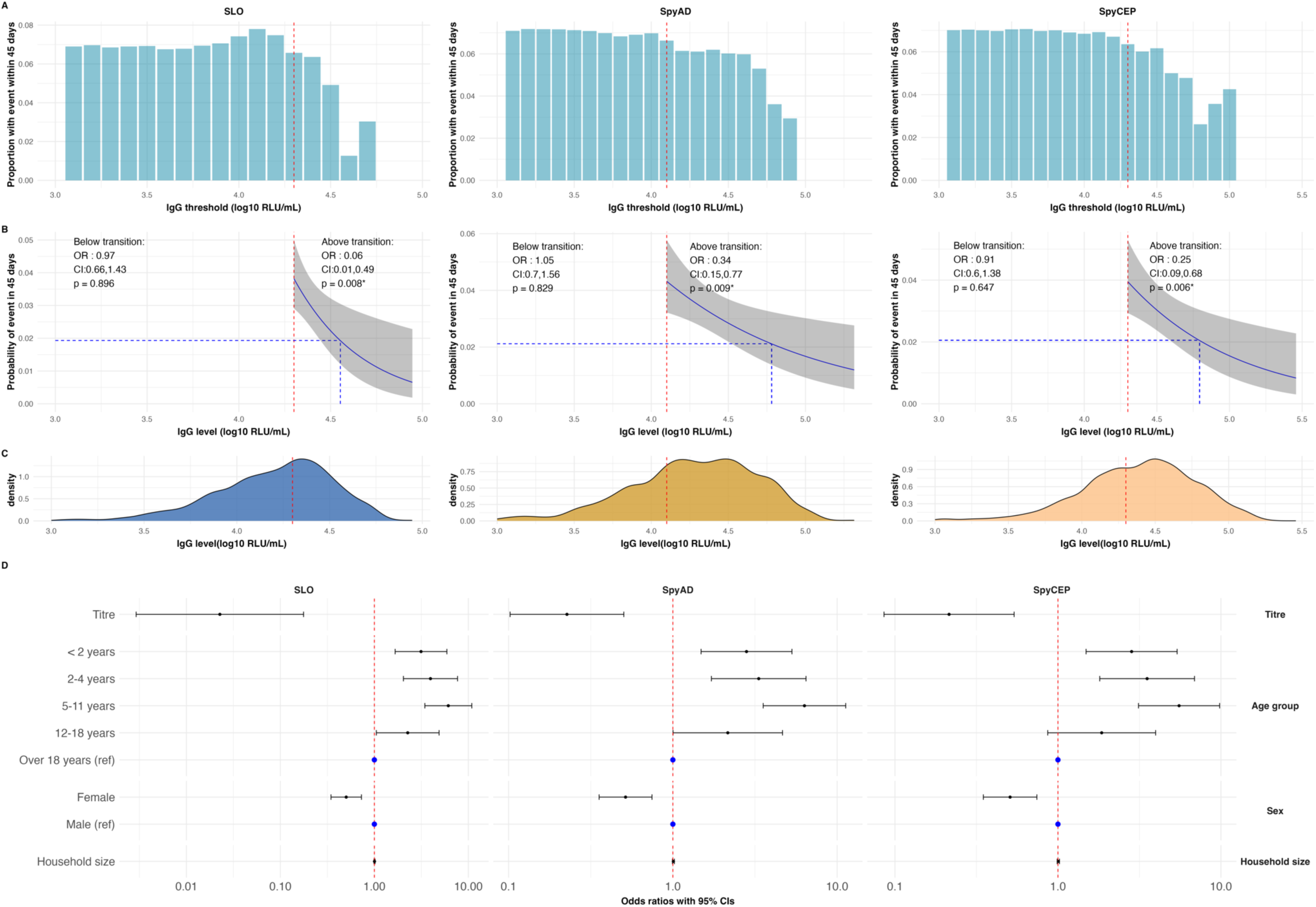
Association between IgG levels to conserved vaccine antigens and protection from culture confirmed S. pyogenes events. (A) The proportion of visits with IgG levels above each threshold associated with a culture-confirmed *S. pyogene*s event within 45 days. IgG levels were measured from n=1,987 visits. (B) Piecewise logistic regression analysis with mixed effects to explore the relationship between IgG level and event within 45 days. Transition points in the relationship between titre levels and event risk were identified from panel A and refined using Akaike Information Criterion (AIC) analysis. Piecewise logistic regression with titres above and below the transition point was performed. The blue line shows the fitted regression model, capturing the association between titre levels above the breakpoint and the probability of a *S. pyogenes* event within 45 days, grey shading represents the 95% confidence intervals for model predictions. Odds ratio and 95% confidence intervals, and p-values for the piecewise model are displayed. The red vertical line indicates the transition point. Putative 50% protective thresholds were calculated as the IgG level at which the predicted probability of an event with 45 days was 50% that of the predicted probability at the transition point. 50% thresholds are plotted with blue dotted lines. (C) Density plot showing the distribution of IgG measurements (n=1987) in realtion to IgG level. The red line marks the transition identified in panel B. (D) Forest plot to visualise the association between IgG level above transition point for each conserved antigen and any culture-confirmed event within 45 days. Odds ratios were calculated from a mixed effects logistic regression model adjusting for factors known to alter risk of *S. pyogene*s events: age, sex and household size. OR = Odds ratio, CI = 95% confidence interval, RLU = Relative Luminex Unit.

To obtain putative 50% protective thresholds for SLO-, SpyAD- and SpyCEP-IgG, an unadjusted mixed-effects logistic regression was used to predict the IgG level at which the probability of any event was 50% of the probability at the transition point (Fig 4B). These thresholds were 35,739 RLU/mL for SLO, 60,345 RLU/mL for SpyAD, and 62,209 RLU/mL for SpyCEP. At study baseline, IgG levels were above these thresholds in 64 individuals (15%) for SLO, 56 (14%) for SpyAD, and 80 (19%) for SpyCEP (Fig 5A).

**Figure 5:**
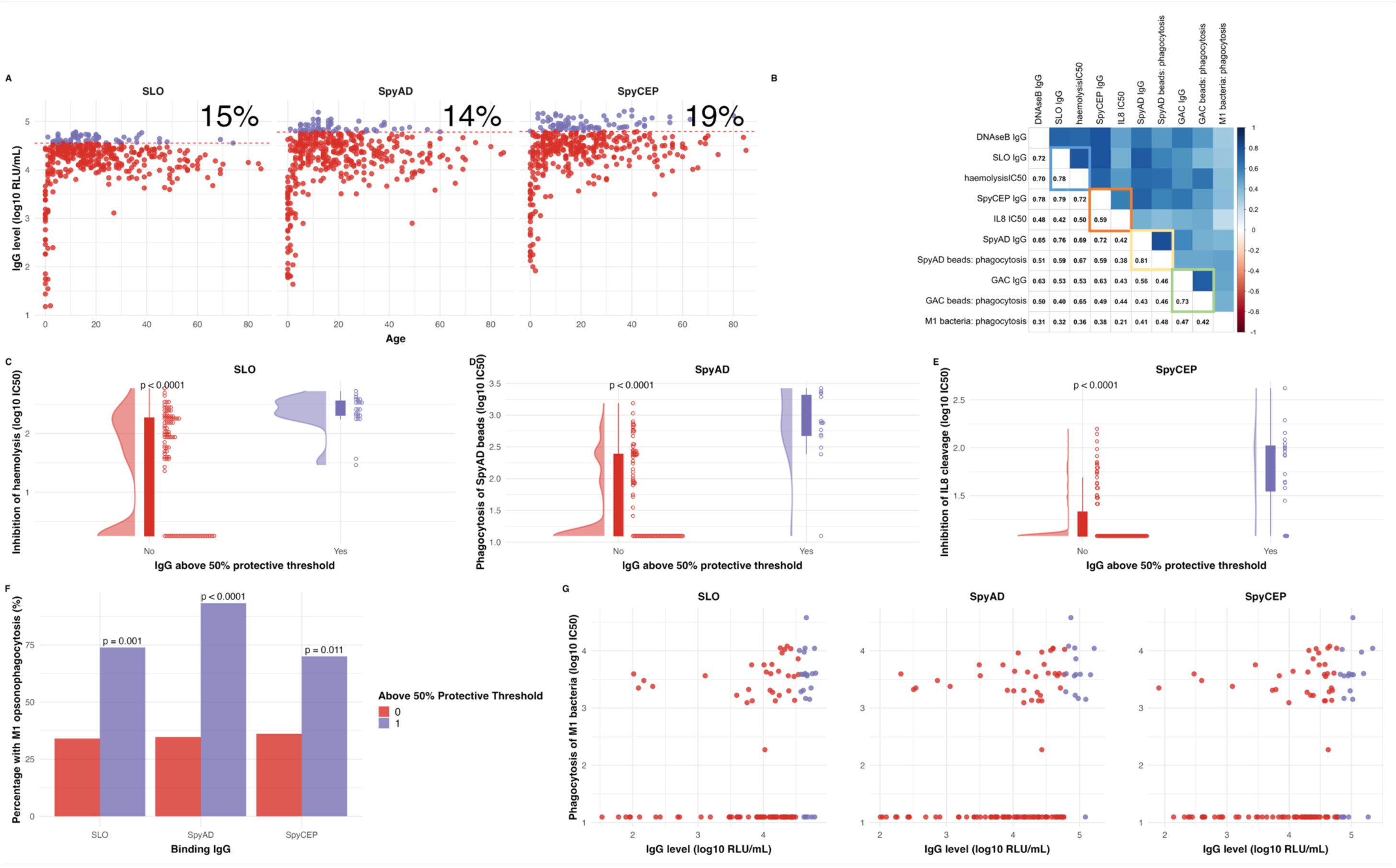
Association between protective IgG profiles and in vitro inhibition of function and promotion of opsonophagocytosis. (A) IgG titre distribution by age at study baseline (n-413), including the percentage of participants with titres above (purple) identified 50% protective threshold. (B) Spearman correlation coefficients between binding IgG titres and functional immunoassays in n=114 serum samples randomly selected across a broad range of binding IgG levels. Highlighted squares indicate the specific relationship between binding titres and the immunoassay that directly measures the function of the corresponding antigen. Blue squares represent SLO, orange represent SpyCEP, yellow represent SpyAD, and green represent GAC. (C-E) Relationship between binding IgG levels and functional activity in serum samples (n=114): (C) IgG binding levels to SLO and inhibition of SLO-mediated hemolysis, (D) IgG binding levels to SpyAD and promotion of phagocytosis of SpyAD-coated beads by THP1 cells and (E) IgG binding levels to SpyCEP and inhibition of SpyCEP-mediated IL-8 cleavage. IC50 between those above and below 50% protective thresholds were compared with Wilcoxon tests. (F) Proportion of participants with titres above and below 50% protective thresholds, with any detectable opsonophagocytosis of M1 bacteria. Proportions between groups were compared with a fisher exact test. (G) Relationship between IgG binding titres to SLO, SpyAD, SpyCEP, and opsonophagocytosis of M1 bacteria by THP1 cells. Binding IgG level above (purple) and below (red) the 50% protective threshold is demonstrated.

To confirm our findings using an orthogonal approach, an Anderson-Gill extension of Cox proportional hazards model was used, as described previously to establish epidemiological risk factors for *S. pyogenes* in this study.^10^ IgG levels above the transition point were added as time dependant covariates, accounting for clustering within households and repeated events in individuals, with adjustment for age group, sex, and household size. Higher IgG levels to SLO (Hazards ratio (HR) 0.04, 0.01-0.23, p=0.00036), SpyAD (HR 0.29,0.16-0.53, p<0.0001) and

SpyCEP (HR 0.38, 0.16-0.90, p=0.027) were associated with protection from any culture-confirmed event (Table S1). In models stratified by event type, protection against *S. pyogenes* carriage events was associated with higher IgG to SLO (HR 0.01, 0.00-0.13,p=0.00012), SpyAD (HR 0.18,0.08-0.41,p<0.0001) and SpyCEP (HR 0.20, 0.09-0.49,p=0.00037; Table S1), which was driven by protection from pharyngeal but not skin carriage (Table S2).

### Protective IgG levels in serum are associated with in vitro functional activity and opsonophagocytosis

To explore the relationship between binding IgG levels and functionality, a sub-sample of 114 sera were randomly selected within IgG strata to represent a wide range of antibody levels. Assays were used to measure the ability of sera to inhibit SLO-mediated haemolysis of erythrocytes,^34^ SpyCEP-mediated interleukin 8 (IL8) cleavage,^35^ and potentiation of THP-1 cell phagocytosis of SpyAD-bound beads. Opsonophagocytic activity against GAC-bound beads and whole M1 *S. pyogenes* was also assessed.^36^

Positive correlations between binding IgG levels and functional activity were strongest for anti-SpyAD (0.81, p<0.0001), followed by SLO (0.78, p<0.0001) and GAC (0.73, p<0.0001), with a modest correlation between anti-SpyCEP IgG and inhibition of IL-8 cleavage activity (0.59, p<0.0001; Fig 5B). Sera with binding IgG levels above putative 50% protective thresholds demonstrated significantly higher functional activity against SLO (p<0.0001; Fig 5C) and SpyCEP (p<0.0001; Fig 5E), and significantly higher opsonophagocytosis of SpyAD-coated beads (p<0.0001; Fig 5D). Opsonophagocytosis of whole M/*emm*1 bacteria was observed in a greater proportion of serum samples with IgG levels above the 50% protective threshold, compared to below it, for SLO (74% vs 34%, p=0.00078), SpyAD (93% vs 35%, p<0.0001), and SpyCEP (70% vs 36%, p=0.011, Fig 5F). Modest but statistically significant correlations were seen between binding IgG levels to SLO (0.32, p<0.00044), SpyAD (0.41, p<0.0001), SpyCEP (0.38, p<0.0001), and opsonophagocytic activity against whole M1 bacteria (Figs 5B & G).

### IgG levels to M/emm cluster representative peptides are more heterogenous than conserved antigen IgG

To compare anti-M humoral immunity with conserved antigen-specific IgG, antibody levels to a range of M peptides were measured. Although over 275 *emm* types exist, several M/*emm* clusters have been defined based on M protein structural similarities. *In vitro* cross-reactivity (and potentially cross-protection) may exist within each cluster.^15,22,25^ In baseline cohort sera, IgG levels to 14 M/*emm ‘*cluster-representative’ 50mer HVR M peptides demonstrated more heterogeneity across the life course than for conserved antigen-specific IgG (Fig 6A).^21^ Raw median fluorescence intensity (MFI) antibody levels (unadjusted to levels in standard material) across all antigens showed a hierarchy of signal ranging from high SLO, SpyAD, SpyCEP, GAC and DNAse B, to moderate levels of anti-M4, anti-M89 and anti-M75 IgG, and lower or heterogenous levels of other M-specific antibodies (Fig 6B). At birth IgG to all M peptides were significantly lower in cord serum than paired serum from mothers (Fig S5).

**Figure 6:**
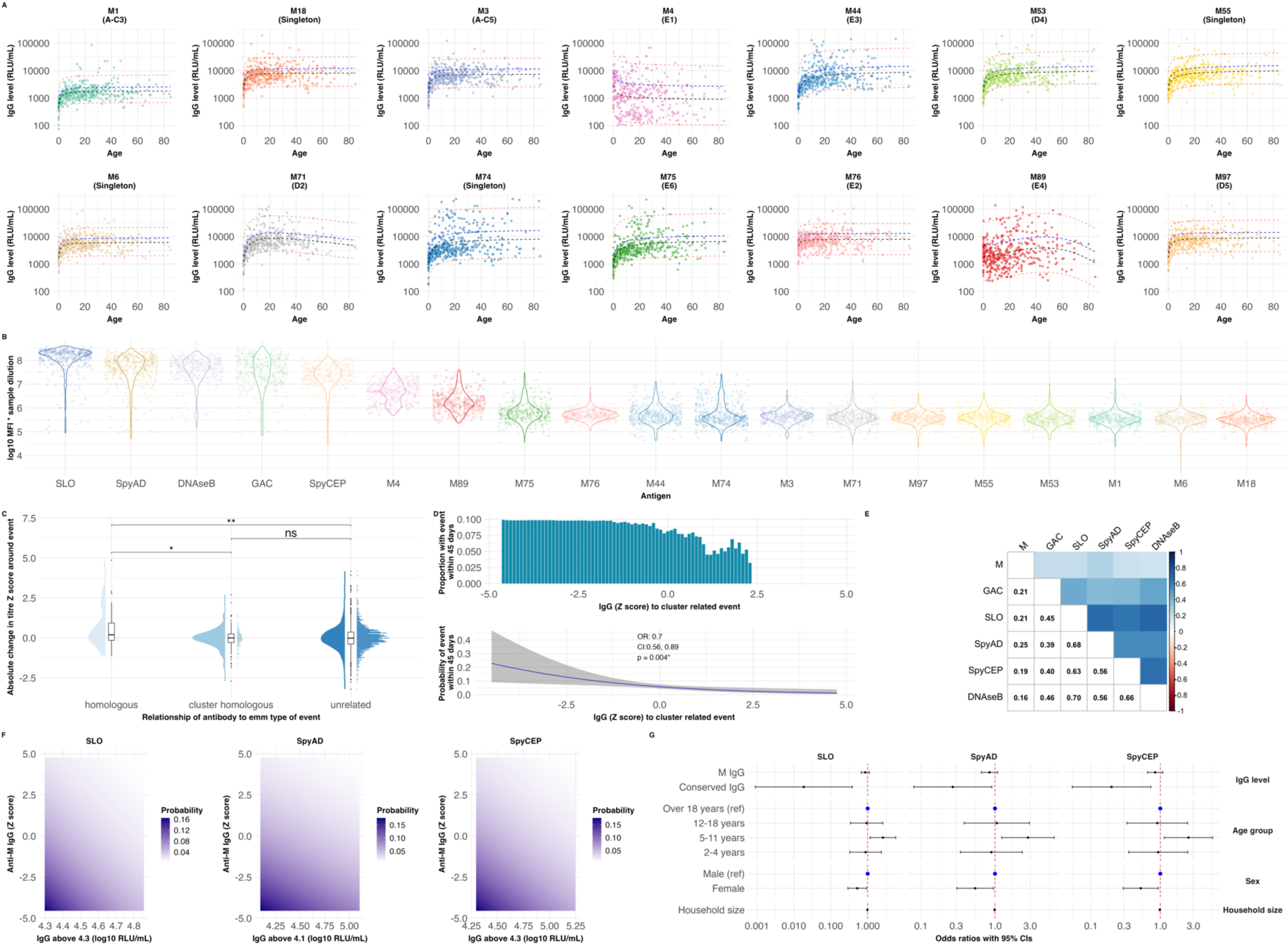
Exploring the role of type specific anti-M protein antibodies in protective immunity. (A): Cross-sectional anti-M IgG profiles in those with no disease events at baseline visit, in sera from the SpyCATS cohort study (n=402) by age. (B): IgG antibody titres at study baseline displayed as raw median fluorescent index (corrected for sample dilution), demonstrating the relative abundance of specific IgG in participant sera. (C): Absolute change in anti-M IgG Z-scores around n=131 culture-confirmed, M/*emm*-typed *S. pyogenes* events. Titres were categorized as: homologous: where the M protein measured was identical to that of the *emm* type in the event (n=40 paired measurements), cluster homologous: the M protein measured was within the same *emm*-cluster(n=201 paired measurements), but not the same *emm* type, as the event, or unrelated: where the M protein measured was not related to the *emm* type of the event(n=1776 paired measurements). The absolute change in Z-score was compared between multiple groups using the Kruskal-Wallis test, followed by Dunn’s test with Bonferroni correction. Significance is indicated as: P < 0.05 (*), P < 0.01 (**). (D): Anti-M IgG before, during and after culture-confirmed M/emm type-characterized events in both cases (n=378 measurements, 143 events, 103 participants) and household controls (n=1360 measurements, from 293 participants) was measured against the *emm*-cluster related M peptide to the M/*emm* type of the event. The cluster-related IgG level was assigned hierarchically with the homologous titre included where available, otherwise the *emm* cluster homologous titre was selected. The association between Z-score normalized anti-M IgG and the occurrence of any event within the next 45 days was explored using logistic mixed effects models. (E): Correlation coefficients (Spearman method) between anti-M Z score normalized IgG levels and the conserved antigen IgG level from 1648 measurements used to explore within *emm-*cluster protection. (F): Mixed effects logistic regression models, including anti-M IgG Z scores to cluster-related and conserved antibody IgG levels above transition point. The density plot represents the probability of an event occurring within 45 days, based on both anti-M, and conserved IgG levels. (G) Odds ratios of an event occurring within 45 day in fully adjusted mixed-effects logistic regression models accounting for anti-M IgG, conserved antigen IgG level (above transition point), age group, sex and household size. OR = Odds ratio, CI = 95% confidence interval, RLU = Relative Luminex Unit.

Each culture-confirmed *S. pyogenes* event was *emm*-typed as previously reported,^10,37^ allowing antibody data in relation to events to be categorised as homologous (M peptide matching the event M/*emm* type), cluster-homologous (non-matching M peptide in same *emm*-cluster as event M/*emm* type), or unrelated. Antibody levels were measured in paired pre/post samples to 14 cluster-representative M peptides and to 5 additional M peptides from the E3 *emm*-cluster, which contains *emm-*types seen commonly in The Gambia (n=2017 paired measurements from 131 *emm*-typed events). Anti-M IgG RLU/mL were Z-score transformed to allow aggregation and comparison across *emm*/M-types. Absolute Z-score increases before and after events were categorised into homologous (M peptide matching the event *emm* type, n=40), cluster-homologous (non-matching M peptide in same cluster as event *emm* type, n=201), or unrelated (n=1776) responses. Absolute increases in IgG levels induced by *S. pyogenes* events were greater for homologous events than for cluster-homologous (p=0.0149) and unrelated events (p=0.0080, Fig 6C).

### IgG levels to conserved vaccine antigens are associated with protection independent of anti-M IgG responses

We next explored whether anti-M IgG was associated with protection. For each emm-typed event (n=143), the homologous or, if unavailable, the cluster-homologous anti-M IgG Z-score (herein called cluster-related) was identified in cases before, during, and after the event, and in household contacts before and after the event. 1649 measurements from 1214 timepoints and 307 individuals were included. For comparison, a mean Z-score of event-unrelated anti-M IgG was also generated for each timepoint. Cluster-related IgG was correlated with mean unrelated anti-M IgG (correlation coefficient 0.59, p<0.0001).

In a mixed-effects logistic regression model, higher cluster-related anti-M IgG was associated with lower odds of any culture-confirmed event (OR 0.7, 95% CI 0.56-0.89, p=0.004, Fig 6D), which was confirmed in a model adjusted for age, sex, and household size (OR 0.75, 0.59-0.97, p=0.025, Table S3). Piecewise regression models for anti-M Z scores demonstrated higher AIC scores. Furthermore, models replacing cluster-related IgG with mean event-unrelated M IgG explained the data less well (AIC 815 vs 804).

Unlike the strong correlation and collinearity *between* conserved antigen IgG (Fig 2D), correlation between cluster-related anti-M IgG and each conserved antigen IgG level was low (coefficients 0.16-0.25, Fig 6E). We therefore sought to establish the relative contribution of conserved and anti-M IgG to protection, using AIC criteria to identify the best fitting model and exclude significant interactions between IgG to M and conserved antigens. IgG levels above transition point for SLO, SpyAD and SpyCEP were included in mixed-effects logistic regression models, along with cluster-related anti-M IgG Z score, age, sex, and household size. Anti-SLO (OR 0.02, 95% CI 0.00-0.38, p=0.010), -SpyAD (OR 0.27, 0.08-0.91, p=0.034), and -SpyCEP (OR 0.20, 0.05-0.73, p=0.0015) IgG were independently associated with protection in each model (Fig 6F&G). An independent but non-significant trend towards protection with cluster-related anti-M IgG was also observed. Of note, the specificity of some anti-M assays was limited, as assessed by competitive inhibition, likely in part due to low median fluorescence intensity (MFI) in IVIG derived from HIC. Despite optimising this signal as best possible with new pooled standards, specificity remained low for some anti-M assays (Fig S6,7). Sensitivity analyses using only seven M peptides with the best specificity demonstrated consistent findings across all M-related analyses (Fig S8).

## Discussion

In a high burden setting for *S. pyogenes* disease, we demonstrate that serological profiles are driven by intense exposure in the first years of life. We also demonstrate, for the first time, that high IgG levels to SLO, SpyCEP and SpyAD are associated with protection from culture-confirmed events, independent of type-specific antibodies. Importantly, these conserved antigens are included in several *S. pyogenes* vaccines in development.^3,26,28,30,38^

After waning of maternal IgG, we observed a rapid rise and plateauing of serum IgG levels in the first few years of life and gradual waning of IgG to some antigens in older adults, similar to findings from Fiji and Uganda.^39,40^ This likely reflects heavy exposure to *S. pyogenes* in this environment, in keeping with the high incidence rates of 409/1000 person-years for carriage and 542/1000 person-years for disease in children under five years in The Gambia.^10^ The most vigorous responses occurred in children under two years, regardless of site or presence of symptoms. This likely explains why low pre-event levels, found most commonly in young infants, were strongly correlated with greater absolute rises in IgG following exposure, consistent with prior observational and human challenge studies^29,41,42^. Immune responses to *S. pyogenes* pharyngitis have been extensively characterized, yet our data demonstrate that pharyngeal carriage, skin disease and skin carriage are equally important drivers of early life immunological responses.^27,29,43,44^ Interestingly, adults with culture-confirmed *S. pyogenes* disease or carriage demonstrated limited boosting of IgG to conserved antigens, highlighting the limitations of using streptococcal serology to provide evidence of recent infection in high-burden settings. It is possible that IgG boosting may be greater during toxin-mediated or invasive *S. pyogenes* infections and in the context of acute rheumatic fever (ARF), which were not explored in our study.^45^ Interestingly, while placental transfer of IgG to conserved protein antigens was complete, transfer of IgG to GAC and M peptides was significantly lower, perhaps explained by previously-documented polarisation of natural anti-GAC responses to IgG2 and anti-M to IgG3,^46^ with differential placental transfer of such isotypes.^47^

Understanding the extent of early life exposure is important in determining the age at which *S. pyogenes* vaccines should be introduced. We observed that 23% of infants have experienced a likely serological priming event by 11 months, yet only 2% of participants under two years had baseline IgG levels above our putative 50% protective threshold for each antigen. Recent modelling assuming protection in line with WHO preferred product characteristics suggested that introduction of a *S. pyogenes* vaccine in five-year-olds would have greater impact than vaccinating at birth.^48^ However, data from New Zealand suggest that exposure to diverse streptococcal genotypes alongside heightened serological responses in early life may be important priming events for ARF and RHD.^43,45^ The intense serological activity we observed in early life may suggest that pathological immune priming in susceptible individuals begins before the age of five. Enhanced protection earlier in life could be critical in preventing RHD in high burden settings. Future vaccine trials and modelling should evaluate the impact of vaccinating children under the age of two years. On the other hand, we observed substantial heterogeneity in IgG levels to conserved vaccine antigens even in adolescence and adulthood. Given only a minority of participants demonstrated levels above our putative 50% protective thresholds, a successful vaccine could boost protective responses even in older children. As the assays described will be used in clinical evaluation of a leading *S. pyogenes* vaccine, our study provides valuable data against which early-phase vaccine trial immunogenicity can be compared. It is crucial, however, that our data are calibrated to international *S. pyogenes* antibody standards when developed in the future, allowing wider comparison and validation.

Future vaccine trial endpoints will likely be *S. pyogenes* pharyngitis and skin infection, not carriage. It is important to note that the endpoints studied here were all *S. pyogenes* events, and it is plausible that the immune responses required to prevent pharyngitis and pyoderma are different to those that protect against carriage. While our study lacked power to explore protection from specific events, especially with low pharyngitis incidence, the clearest protective signal was observed to pharyngeal carriage, despite similar numbers of skin carriage and disease events. Environmental factors such as physical trauma and hygiene may be more important in the pathogenesis of *S. pyogenes* skin events, whereas humoral immunity may play a greater role in preventing pharyngeal events.^10,49^ This has implications for evaluating whether vaccine-induced immunity can prevent pharyngitis and pyoderma in future trials.

While our conserved antigen IgG assay has been extensively characterized,^32^ our anti-M IgG assays suffered from poor specificity for some peptides. This may be due to a combination of anti-M cross-reactivity and variable amounts of type-specific IgG in the IVIG/pooled serum used for assay validation. Despite limitations, we observed low and heterogenous anti-M IgG levels at study entry to multiple type-specific HVR peptides, with the highest responses seen to M peptides from *S. pyogenes* types observed more frequently in The Gambia, including *emm*4, *emm*89 and *emm*75.^50,51^ The streptococcal M protein is a major immunogen and potential vaccine target, with type specific antibodies shown to be protective *in vitro* and in limited observational data.^15,17–19,24,52^ In contrast to conserved streptococcal antigens, type-specific IgG likely accumulates more slowly in settings like The Gambia with high *emm* diversity.^50,51,53^ At present, multivalent vaccines to M protein HVR, based largely on *emm*/M types common in HIC, are the only *S. pyogenes* vaccines to have progressed to phase II trials.^54^ The efficacy of multivalent M-based vaccines would rely on the degree of cross protective immunity within M/*emm* clusters.^21,22,25^ A protective effect of cluster-related anti-M IgG was apparent, although these analyses were likely limited by power.

Our findings that IgG levels to conserved antigens SLO, SpyAD and SpyCEP may be associated with protection, independent of anti-M immunity, is particularly pertinent to LMICs where M/*emm* type diversity is greatest.^20,21^ We demonstrate a strong correlation between binding IgG and functionality, though SpyCEP-mediated IL-8 cleavage inhibition was less clearly correlated with anti-SpyCEP IgG, perhaps explained by binding of antibodies to non-functional regions of SpyCEP. The association between IgG binding levels and opsonophagocytosis of *emm*1 bacteria is also likely mediated by antibodies to conserved targets, either measured or unmeasured, given low levels of anti-M1 IgG in the cohort and no prior documentation of *emm*1 bacteria in The Gambia in multiple studies.^37,50,51^ Of note, higher IgG to GAC and DNAseB were not associated with protection in our data. GAC is a promising preclinical vaccine candidate contained within several leading vaccines and vaccine-induced protection may differ significantly from naturally-acquired immunity.^3,55–58^ Importantly, the anti-SLO, -SpyAD and -SpyCEP IgG associated protection we observed may also be a surrogate of unmeasured adaptive immune responses. Future vaccine trials and human challenge studies will help establish whether these are true mechanistic correlates of protection.^7^

Our study has additional limitations. We likely missed *S. pyogenes* events for several reasons. We have previously demonstrated the limited sensitivity of culture compared to molecular methods in this setting.^53,59^ Nonetheless, culture positivity is directly related to quantitative PCR bacterial load,^53,59^ and therefore remains a relevant, if insensitive, outcome. Secondly, our monthly routine sampling likely missed carriage events, given the short carriage duration of *S. pyogenes* (median 4 days) we have reported.^10^ The immunological responses observed in children under two years with no culture-confirmed events reflects this. The framework used to select timepoints for antibody measurement and assessing protection, rather than unbiased universal testing of all samples, may have introduced biases. We mitigate this by demonstrating that IgG levels in older participants without culture-confirmed events remained broadly constant and by testing every timepoint in participants under the age of two. Furthermore, we tested all samples from culture-confirmed cases and household contacts longitudinally around events, where antibody levels were most likely to change.

In conclusion, our study represents a unique resolution of immunological sampling from intensively-followed cohorts across the life course in a high burden setting for *S. pyogenes* and RHD, using robust and reproducible immunoassays,^32,34–36^ focusing on antigens within leading candidate-vaccines.^3^ We demonstrate the dynamic evolution of humoral immune responses in children under two years, a previously underrepresented group in observational studies. Our data suggest that antibodies to both conserved vaccine antigens and M peptides may be associated with protection from culture-confirmed *S. pyogenes* events, providing optimism for both conserved and multivalent M-protein approaches. Further well-designed epidemiological and multiphase clinical vaccine trials in high burden settings are urgently required to further understand mechanisms of protective immunity and to identify a tractable correlate of protection.

## Materials and Methods

### Study Participants and sampling

Mother child pairs during first year of life: Mother child pairs from an urban clinic, The Gambia, participating in a trial of maternal immunisation with MenAfriVAC were included (NCT03746665). All samples were obtained from consenting mother-child pairs where the mother had been vaccinated with meningococcal serogroup A conjugate vaccine between 28 - 34 weeks’ gestation. Participants were vaccinated between December 2018 and October 2019. All participants were included where a paired serum sample from mother at delivery and neonatal cord blood was available.

Household longitudinal cohort study: Participants in the *Streptococcus pyogenes* carriage acquisition, persistence and transmission dynamics within households in The Gambia (SpyCATS) household cohort study were included.^10,60^ A total of 442 participants from 44 households were recruited and visited monthly. At each monthly visit, all enrolled participants were swabbed to determine the presence of Group A beta-hemolytic streptococci (GABHS) in their normal skin and throat. Unscheduled visits took place at any time when participants reported skin sores or a sore throat (disease episodes) to the study team. Suspected disease sites were swabbed for GABHS. At baseline, a dried blood spot (DBS) was taken from all children and a blood sample for serum separation from all children over 2 years of age. A DBS was collected from all participants during each monthly visit, and at any presentation with disease.

### Microbiologically confirmed event definitions from SpyCATS longitudinal cohort study

Disease events were defined as the presence of signs or symptoms of pharyngitis or pyoderma plus a positive culture for *S. pyogenes* from the disease site. Carriage events were defined as the detection of *S. pyogenes* from throat or skin swabs without symptoms or signs of disease. We categorised events into two distinct types: Response-Focused Events (RFE) and Protection-Focused Events (PFE). RFEs were defined to study immune responses to specific events. RFE carriage events could only be defined in absence of a disease event with preceding 42 or following 14 days. PFEs were defined as previously, and used to analyze risk factors for incident events, excluding weekly swabs from carriage incidence analysis, and allowing simultaneous characterisation of disease and carriage events.^10^ A detailed description and breakdown of event characterisation is provided in supplementary methods.

### Sample selection for IgG level measurement

For mother child pairs, samples used were those from mothers at delivery, neonatal cord blood at delivery, and samples from infants taken at 6 months, and from between 9- and 11-months of age. From the SpyCATS cohort, a baseline sample of serum (or DBS in under two-year-olds) was selected from all participants. DBS samples selected around RFEs were taken from the closest sample 14 days prior to the event (pre-event), the time of the event (event), and the closest sample at least 14 days after the event (post-event), as well as 3 and 6 months after the event where available. We identified a control group of exposed, uninfected household contacts. These individuals were in the same household as an index case, were sampled within 14 days of the index event, had no *S. pyogenes* event within 90 days of the index case, and had a DBS sample taken on the day of the household event or within the 28 days prior. The control group also had a sample selected for testing between 14 and 42 days from the index event where available. For the cross-sectional analysis of age-stratified IgG levels, a single measurement per participant was taken from baseline, providing there was no *S. pyogenes* event at the time. In the case that both serum and DBS were measured at a single timepoint, a geometric mean IgG level was taken from the two readings. Additionally in participants under the age of two years, where IgG levels were rising fastest, a DBS sample was tested from every time point collected in the study.

### IgG measurement in blood for conserved S. pyogenes antigens

Both serum and DBS were tested with a Luminex 5-Plex assay to measure IgG levels to the conserved *S. pyogenes* antigens GAC, SLO, SpyAD, SpyCEP and DNAseB, using methods previously described.^32,33^ Very strong correlation of serum with eluted DBS was established (Supplementary methods, Fig S9).^33^ The Luminex assay was conducted as previously described with sample dilutions ranging from 1:300 to 1:60,000 and beads added at 1000 beads/region/well. A standard curve of Privigen intra-venous immunoglobulin (IVIG) in three-fold dilutions from 1:990 was added to each plate alongside a single sample of pooled serum to act as a positive control. Samples were tested in single, at starting concentration of 1:20000, and retested at an alternative dilution if the mean fluorescence intensity (MFI) value fell outside of limits of standard curve accuracy.^32^ All serial DBS samples from the same individual were tested on the same assay plate. The geometric mean level from all repeats falling within limits of standard curve accuracy was taken as the final level.

### IgG measurement in blood for type-specific M hypervariable region peptide antigens

We further adapted the Luminex assay to measure IgG to 14 M*/emm* type specific M peptides, selected as representative of different M/*emm* clusters,^21^ along with five additional M peptides from the E3 M/*emm* cluster, which were most common in the study (Table S4).^10^ Biotinylated 50-amino-acid peptides derived from the N-terminal hypervariable region (HVR) of each M/*emm* type, were coupled to MagPlex beads, pre coupled to Streptavidin as described.^32^ Bead coupling concentration of M peptides was 5μg per million beads. The assay was optimized to measure M-protein antibodies at 1:2500 dilution of samples. Cluster-representative 14-plex coupled beads were added to the diluted samples at 1000 beads/region/well in 10μL. Standard material consisted of 25% IVIG (Gammanorm, Octagen), 25% pooled sera from n=9 participants who experienced documented *emm*25, *emm*18 and *emm*113 events, and 50% pooled human sera from SpyCATS study final visit (n=244). This combination was selected to enhance type-specific antibody detection for M peptides with low MFI in commercial IVIG preparations (collected in high income countries). Assay specificity was characterized (Supplementary methods, Fig S5,6). Each assay plate included a 10-step serial dilution of the standard, starting from 1:100. The same incubation conditions were applied as previously described.^32,33^ An additional 6-plex assay containing 6-peptides from the E3 cluster was also employed on specific samples to investigate responses to E3-related events, under the same assay conditions as for cluster-representative M-peptide IgG measurement. The cluster representative 14-plex assay was performed on every sample selected for measurement in the study. The additional E3 6-plex assessment was performed only around events from the E3 *emm* cluster.

### Functional immunoassays

The inhibition of streptolysin O (SLO)-induced haemolysis by sera was assessed using a previously characterized assay.^34^ Serial dilutions of sera were incubated with SLO and red blood cells, and after incubation, haemolysis was measured by absorbance at 540 nm. The IC50 value was determined, representing the serum concentration required to inhibit 50% of haemolysis.

To measure the inhibition of SpyCEP-mediated IL-8 cleavage, sera were incubated with enzymatically active SpyCEP and IL-8, and IL-8 levels were quantified using a sandwich ELISA as previously described in detail.^35^ The IC50 was calculated as the serum concentration required to achieve 50% inhibition of IL-8 cleavage. Opsonophagocytic activity was evaluated using a previously described assay.^36^ where FITC-labelled bacteria and antigen-coupled beads (SpyAD and GAC) were incubated with sera and THP-1 cells. Phagocytosis was assessed by flow cytometry, measuring the geometric mean FITC intensity within the THP-1 cell gate. IC50 values were calculated representing serum dilution at which 50% of maximum phagocytic activity was observed. For detailed methods see supplementary data.

### Statistical analyses

To obtain relative IgG levels, a five-parameter logistic (5PL) curve was fit to the blank-subtracted MFI values obtained for each standard curve point using Bioplex manager software. Relative Luminex Units per milliliter (RLU/mL) for each dilution of test samples were obtained by interpolation of the blank-subtracted MFI values at each specific dilution to a 5PL curve, multiplying this value by the dilution factor. RLU/mL values were log10 transformed for statistical analysis. IC50 data during functional assays were produced in Graphpad (v10), by fitting four-parameter logistic (4PL) curves to data. In functional assay analyses, samples with an IC50 below the limit of quantification (LOQ) were assigned an IC50 value of half the LOQ as described in assay characterisation.^34–36^ All remaining statistical analysis was performed with R (version 4.4.0). Fractional polynomial models were applied to log10 transformed cross-sectional antibody data to determine the 2.5%, 50%, 80% and 97.5^th^ centile for each antigen by age, using previously described methodology.^40,61^

After assessment of both IgG levels and absolute antibody level changes around RFEs with QQ-plots, histograms and the Shapiro-Wilk test, non-parametric tests were used. After additional testing for homoscedasticity by plotting residuals, the Pearson method was used to determine correlation coefficient with levels and absolute level changes within individuals, given large sample sizes. Functional immunoassay data and IgG levels to M peptides were assessed for correlation with Spearman method. When IgG levels were compared between two groups a Mann Whitney U test was used, corrected for testing across multiple antigens using false discovery rate (FDR) method. For multiple comparisons between groups, Kruskall-Wallis followed by Dunn’s test with Bonferroni correction was used. Exploring the impact of age and event type on magnitude of absolute log10 IgG changes around events was done with a mixed-effects linear regression analysis accounting for individuals and households as random effects. P values (adjusted where appropriate) below 0.05 were considered significant.

Protection associated with of IgG level and culture confirmed events (PFEs) was explored using both logistic regression models and the Anderson Gill extension of Cox proportional hazards models.^10,62^ IgG measurements included in analysis of protection from conserved antigens were from all baseline samples, all samples in participants aged under two years, samples before, during and after microbiologically confirmed *S. pyogenes* events in cases, and samples before and after a microbiologically confirmed events in household controls. IgG levels were assumed to remain constant between measurements. Mixed-effects logistic regression models were used to explore the association of IgG with an event at the next visit so long as the next visit occurred within 45 days, to account for the monthly sampling frame in the SpyCATS study. Random effects for individuals and households were used to account for repeated sampling from individuals and household structure within the study. P values below 0.05 were considered significant. Models were constructed separately for each antigen given substantial collinearity between conserved antigens. To construct piecewise regression for protection mediated by SLO, SpyAD and SpyCEP, the transition point at which proportion of events above each IgG threshold began to diminish were identified visually for each antigen. Next the AIC values for, 0.1 log10 iterations of the transition point were compared to the AIC of non-piecewise logistic regression, ensuring AIC values were at least 2 lower using piecewise regression, and to confirm the appropriate transition point. AIC values <2 were considered comparable. In adjusted models, AIC values were used to choose the model to best explain the data and to justify the inclusion of level above the transition point only. Final models selected through this method explored the association between event in the next 45 days and IgG level above transition point, sex, age group and household size. To establish 50% protective threshold for IgG levels in blood to SLO, SpyAD and SpyCEP, the IgG level at which the probability of any event in the next 45 days was 50% compared to IgG level at the transition point.

To explore association between IgG levels to conserved antigens and protection using an orthogonal approach, the Andersen-Gill extension of the Cox proportional hazards model was used to explore the association of antibody level with incident events, as described.^10^ Outcomes explored in this model included any incident culture-confirmed event, as well as each PFE event category as described. Multivariable models were selected through AIC criteria assessment, including sex, age group, and household size as covariates.

For analysis of M/*emm* cluster protection, each culture confirmed *S. pyogenes* event was M/*emm* typed.^10^ Having identified the cases and household controls, we used the M/*emm* type of the event to allocate a cluster-related IgG Z-score to both cases and controls in relation to each event. If a homologous M peptide measurement (matching the event M/*emm* type) was available, it was selected; otherwise, the cluster-homologous M peptide level (representing the M/*emm* cluster) was used. This measurement was defined as the cluster-related anti-M IgG Z-score. The mean Z score to unrelated M peptides to each event was identified for each timepoint and compared to the cluster-related Z score with Pearson’s correlation. Models incorporating the composite unrelated Z score were compared to those incorporating the cluster-related Z score using AIC criteria. Cluster-related anti-M IgG from before, during, and after the event in cases, and before and after the event in household controls was used to assess for protection from any microbiological confirmed event in mixed-effects logistic regression models as described.

Finally using AIC criteria to select the best model, a mixed-effects logistic regression model was used to explore the association between any event in the next 45 days and IgG level above transition point for conserved antigens, cluster-homologous anti-M IgG Z score, sex, age group and household size

## Data Availability

All code to perform our analyses is currently publicly available and upon acceptance we will release de-identified primary data into the public domain as an open-access resource for the community.

https://github.com/Clinical-Infection-Research-UoSheffield/Development_natural_protective_immunity_Streptococcus_pyogenes

## Code availability

The code and anonymised data to reproduce analyses is available from https://github.com/Clinical-Infection-Research-UoSheffield/Development_natural_protective_immunity_Streptococcus_pyogenes

## Ethical approval

The studies received approval from the joint ethics committee of The Gambia Government/Medical Research Council and the London School of Hygiene & Tropical Medicine Research Ethics Committee (ref: 24005 and 1585). Written informed consent was obtained from adult participants, as well as from parents or guardians for participants under 18 years of age.

Additionally, children aged 12 to 17 years provided assent. The studies are registered on ClinicalTrials.gov (NCT05117528 and NCT03746665).

## Acknowledgements

We thank the Medical Research Council (MRC) Unit The Gambia (MRCG) including the Clinical Services Department led by Dr. Karen Forrest for overseeing the clinical care of study participants, the Research Support Office especially Njilan Johnson, Jebel Cessay, Asheme Mahmoud and Sheikh Omar Jallow, and the Immunology and Microbiology Platforms especially Jainaba Njie-Jobe, Saffiatou Darboe and Martin Goodier; GSK Vaccine Institute of Global Health, for provision of SpyCEP, SLO, GAC, and SpyAD antigens for measuring IgG in Luminex 4-plex assay; Tom Parks at Imperial College London for support with fitting polynomial models to age stratified antibody data and Natalie Lorenz at University of Aukland for advice on M peptide Luminex methodology. Most importantly we thank the study participants and their parents who took part in the studies.

## Contributions

Conceptualization: AJK, FEC, EA, GdeC, PRS, BK, MM, CET & TIdS

Data curation: AJK, FEC, EA, GdeC, MJ, AW, AB, HC, IC, BS, MM

Formal analysis: AJK, FEC, EA, AK, EC, HS, MM, CET, TIdS

Funding acquisition: EA, AJK, AK, EC, MM, BK, GdeC, AB, PRS, CET & TIdS

Investigation: AJK, FEC, EA, GdeC, JS, MLF, VR, ES, MJ, AW, AB, HC, IC, BS, MM, MC, LR, EB, LM, CS, OC, EC, YJJ

Methodology: AJK, FEC, EA, GdeC, MM, MC, LR, EB, LM, CS, OC, EC, MI, DMG, AK, PRS, AB, YJJ, NM, EC, BK, MM, OR, HS, CET & TIdS

Project administration: AJK, FEC, EA, GdeC, MJ, AW, AB, HC, IC, BS, MM, EC, OR, HS, CET & TIdS

Resources: AW, MI, DMG, OR.

Writing: AJK, FEC, EA, GdeC, JS, MLF, VR, ES, MJ, AW, AB, HC, IC, BS, MM, MC, LR, EB, LM, CS, OC, EC, MI, DMG, AK, PRS, AB, YJJ, NM, EC, BK, MM, OR, HS, CET & TIdS.

Functional immunoassays were performed at GSK laboratories by AJK MC EB LR LM. GSK provided antigens and technical support transferring Luminex assay to MRCG in The Gambia.

